# Zero-Dose in Pakistan: A Decadal Investigation of the Roots of Incomplete Immunization

**DOI:** 10.64898/2026.11.27.26369124

**Authors:** Zia-Un- Nisa, Adnan Ahmad Khan, Areesh Fatmee, Olan Naz, Amna Mahnoor Cheema, Muhammad Ibrahim

## Abstract

**Background:** Zero-dose children represent a critical marker of inequity in immunization systems. Despite improvements in national coverage, Pakistan continues to show substantial subnational disparities, particularly in underserved and conflict-affected areas.

**Methods:** We conducted a repeated cross-sectional secondary analysis of Pakistan Social and Living Standards Measurement surveys from 2008–09 to 2019–20 among children aged 12–23 months. Operational zero-dose (OZD) was defined as non-receipt of the first DPT/pentavalent dose, and true zero-dose (TZD) as receipt of no routine vaccines. Provincial trends, district-level choropleth and bivariate vulnerability maps, healthcare-utilization patterns, and multivariable logistic regression were used to examine geographic, socioeconomic, maternal, and service-related correlates of zero-dose status.

**Results:** OZD declined from 25.7% to 18.4% in Sindh, 18.3% to 11.5% in Khyber Pakhtunkhwa, and 49.8% to 36.7% in Balochistan, while remaining comparatively low in Punjab (5.2% in 2019–20). Persistent high-burden districts overlapped with socioeconomic and healthcare vulnerability. In the reported multivariable model, the richest households had lower odds of OZD (adjusted OR [aOR] 0.694, 95% CI 0.689–0.698) and TZD (aOR 0.941, 95% CI 0.931–0.951), and higher maternal education was associated with substantially lower odds (OZD aOR 0.141, 95% CI 0.139–0.143; TZD aOR 0.306, 95% CI 0.300–0.312). Reported lack of cleanliness was strongly associated with OZD (aOR 8.554, 95% CI 7.704–9.499) and TZD (aOR 30.117, 95% CI 27.144–33.416), while uncooperative staff were associated with higher OZD odds (aOR 2.778, 95% CI 2.649–2.913).

**Conclusion:** Although Pakistan has made progress in reducing zero-dose prevalence, substantial geographic and socioeconomic inequities persist. Findings suggest that improving immunization coverage requires not only expanding service access but also addressing underlying vulnerabilities and service quality gaps. Targeted, equity-focused strategies—particularly in high-burden districts—are essential to reach underserved populations and achieve national and global immunization goals.

## Introduction

Vaccine-preventable diseases are a significant cause of morbidity and mortality among children under the age of five due to gaps in immunization coverage worldwide [1]. Investing in immunization in the world’s lowest-income countries yields substantial returns, with each dollar generating an estimated direct return of USD 20, or USD 52 when considering broader societal impacts [2]. Despite advancements made between 2000 and 2019, including the introduction of new vaccines in low- and middle-income countries, many countries still do not meet the World Health Organization’s (WHO) coverage target of 90% [3]. In 2019, 14 million infants were classified as zero-dose [4], with 50% in Nigeria, India, the Democratic Republic of Congo, Pakistan, Ethiopia, and the Philippines. Aligning with the Sustainable Development Goals and Immunization Agenda 2030, Gavi aims to reduce zero-dose children by 25% by 2025 and 50% by 2030 [5].

Zero-dose (ZD) children are not merely under-vaccinated, they are entirely missed by the health systems, reflecting deep rooted inequities. This status signals heightened public health vulnerability due to gaps in herd immunity, as well as systemic failures in governance, financing, and service delivery. To address these structural challenges, Gavi introduced the IRMMA framework (Identify, Reach, Monitor, Measure, and Advocate), which emphasizes identifying zero-dose populations and tailoring interventions to reach underserved communities [6]. In Pakistan, these challenges are evident. According to the Pakistan Social and Living Standards Measurement (PSLM) (2019-20), full immunization coverage was 81% nationally, with substantial provincial variation: Punjab (89%), Sindh (73%), Khyber Pakhtunkhwa (77%), and Balochistan (53%). Despite more than four decades since the launch of the Expanded Programme on Immunization (EPI), Pakistan continues to face persistent immunization gaps at both provincial and district levels. Despite these documented disparities, there is limited longitudinal and district-level evidence in Pakistan that integrates spatial patterns with underlying socioeconomic and healthcare vulnerabilities. This gap restricts the ability to design targeted, equity-driven interventions.

In Pakistan, disparities in immunization coverage are driven by both systemic inequities and missed opportunities. Structural constraints—including limited vaccine financing and unequal service delivery—interact with contextual factors such as socioeconomic insecurity, political instability, and demographic variation [7]. These disparities are reflected geographically: Balochistan’s challenging terrain and Karachi’s urban slums report some of the lowest coverage levels due to systemic exclusion [8], whereas regions such as South Punjab demonstrate comparatively higher uptake, partly attributed to greater parental awareness and positive attitudes toward vaccination [9]. However, existing evidence remains fragmented and does not systematically examine how these structural and contextual factors interact across districts over time.

Zero-dose children in Pakistan are disproportionately concentrated in underserved settings, including remote rural areas, urban slums, and conflict-affected regions [10], where systemic exclusion is most pronounced. For example, the 2022 floods disrupted health service delivery, reducing immunization rates in Sindh by 19% and resulting in approximately 15,000 missed vaccinations [11]. Although targeted interventions in conflict-affected districts such as Lakki Marwat, Peshawar, and Quetta have demonstrated success in reducing zero-dose prevalence [12], scaling such approaches requires granular, district-level evidence on risk factors and microgeographic variation. However, such evidence remains limited in Pakistan.

While some studies conducted in Sindh have explored determinants of immunization coverage [8], other regions remain underexamined. This study addresses this gap by conducting a decadal analysis of PSLM data (2008–2020) to: (1) identify spatial and temporal clusters of zero-dose children at the district level, (2) examine their association with socioeconomic and healthcare vulnerabilities, and (3) differentiate between operational and true zero-dose populations. By integrating spatial analysis with vulnerability profiling, this study provides novel evidence to inform targeted, equity-driven immunization strategies in Pakistan.

## Materials and Methods

### Study design and data source

This repeated cross-sectional observational study used secondary data from the Pakistan Social and Living Standards Measurement (PSLM) surveys conducted in 2008–09, 2010–11, 2012–13, 2014–15, and 2019–20. These surveys provide district-level representation and span more than a decade. Although PSLM is typically conducted biennially, a four-year gap occurred between the 2014–15 and 2019–20 rounds. The surveys include socioeconomic and health indicators suitable for assessing immunization patterns. Data cleaning and statistical analyses were conducted using Stata/SE 17 (StataCorp, USA). Spatial analyses and mapping were performed in Stata using the spmap command after district-level survey estimates were merged with administrative boundary shapefiles.

### Study population and outcome measures

The study population comprised children aged 12–23 months, representing the youngest cohort eligible to have completed the routine infant immunization schedule under Pakistan’s Expanded Programme on Immunization (EPI). This cohort reflects immunization system performance during the first year of life.

#### Eligibility and study size

Children aged 12–23 months with information required to classify vaccination status were eligible for the relevant analyses. Because this was a secondary analysis of existing nationally representative survey rounds, no priori sample-size calculation was undertaken; the analysis used eligible observations available in the selected survey data.

Two key outcome measures were defined:

- ***Operational zero dose (OZD):*** Children who did not receive the first dose of diphtheria– tetanus–pertussis (DPT) or pentavalent vaccine, coded as 1 (not received) and 0 (received). This measure captures initial contact with the health system and is widely used as a proxy for zero-dose status aligned with Immunization Agenda 2030 definitions [2]. ***True zero dose (TZD):*** Children who did not receive any routine vaccines (BCG, polio [OPV/IPV], pentavalent, PCV, rotavirus, or measles), coded as 1 if no vaccines were received and 0 otherwise. This provides a conservative estimate of complete exclusion from immunization services.

Vaccination status was determined using both caregiver recall and immunization records. Using both OZD and TZD allows a more comprehensive assessment of immunization gaps. While OZD reflects lack of initial contact with immunization services, TZD represents complete exclusion from the health system, allowing differentiation between access failure and systemic exclusion.

The routine EPI schedule of Pakistan is detailed in Table 1.

**Table 1.** EPI schedule for pentavalent and measles vaccination in Pakistan.

| Vaccine | Doses | Age of child |
|---|---|---|
| Pentavalent vaccine<br>(DPT+ Hib +Hep. B) | 3 | Penta-1 at 6 weeks |
|  |  | Penta-2 at 10 weeks |
|  |  | Penta-3 at 14 weeks |
| Measles | 2 | Measles-1 at 9 months (36-40 weeks) |
|  |  | Measles-2 at 12 months (52 weeks) |

### Covariates

Household and maternal characteristics associated with immunization uptake were included based on prior literature. Household wealth quintiles were constructed using the Demographic and Health Survey (DHS) asset-based index methodology (principal component analysis), categorized from poorest (Q1) to richest (Q5).

Maternal factors included education level, place of delivery, antenatal care (ANC) visits, and receipt of tetanus toxoid (TT) injections. Child-level healthcare access variables included recent illness (diarrhea), care-seeking behavior, type of facility accessed, and utilization of basic health units (BHUs). These composite measures were constructed based on prior evidence linking socioeconomic disadvantage and maternal healthcare utilization to immunization outcomes [13, 14].

Two composite vulnerability measures were constructed:

1. **Overall vulnerability**, capturing poorest households with no formal maternal education and no ANC visits. Examining overall vulnerability is crucial because economically disadvantaged households with low maternal education levels are less likely to have awareness of and access to essential health services, including childhood immunization, which leads to poorer health outcomes for children (16).
2. **Healthcare vulnerability**, assessing maternal healthcare utilization patterns, such as no ANC visits, no TT injections, and home delivery during the last pregnancy. Similarly, assessing healthcare vulnerability is imperative, as mothers who do not utilize healthcare services, such as antenatal care and TT injections, may miss vital opportunities to obtain information and access to immunization services necessary for their children’s health [14]. Both indices were constructed using a binary aggregation approach, where households meeting at least one of the specified conditions were classified as vulnerable. Sensitivity to alternative definitions (e.g., requiring multiple conditions) was considered in interpretation.

### Statistical Analysis

Descriptive temporal analyses quantified estimates of OZD and TZD by province, adjusted for survey weights, to track changes over time, assess intervention effectiveness, and identify gaps in immunization coverage. Spatial patterns were visualized using univariate choropleth maps, stratified by district across five time periods. Spatial distributions were generated using district-level prevalence estimates and visualized through choropleth mapping. These maps were created by categorizing both variables into predefined classes and overlaying them to visualize their joint spatial distribution and identify co-occurring high-risk districts.

#### Vulnerability Analysis

To identify households facing elevated disadvantage, bi-variate choropleth maps were used to track trends among the most vulnerable population subsets, utilizing overall and healthcare vulnerability indicators in relation to OZD and TZD. For TZD, the cut points established were 0, 0-10, and 10-100, while for OZD, the cut points were 0-10, 10-30, and 30-100. Overall vulnerability was categorized using cut points of 0-5, 5-15, and above 15, whereas healthcare vulnerability was classified with cut points of 0-2, 2-4, and above 4. These cut points were applied to construct the legend shading on the bivariate maps, facilitating clear visualization of vulnerability trends. Cut-points were selected to reflect meaningful programmatic thresholds and to enhance interpretability of spatial clustering, consistent with prior immunization mapping studies.

#### Benchmark Assessment

To pinpoint persistently high-risk districts warranting immediate interventions, the benchmark assessment established thresholds of 50% for OZD and 10% for TZD, in line with the GAVI 5.0 strategy and IA2030 goals, which aim to reduce zero-dose children by 50% and achieve 90% national vaccine coverage, respectively [5]Click or tap here to enter text.. Districts that consistently exceeded these thresholds - specifically, those surpassing them at least three times - were scrutinized for further analysis. Additionally, to ascertain if the shortlisted districts exhibited ongoing trends of suboptimal vaccination coverage and healthcare conditions beyond the study period (2019-2020), government reports on polio cases in districts during 2019-2024 [15] were cross-referenced, enabling an examination of temporal consistency in these trends.

The following analyses, focused on OZD children to determine if their vaccination status results from limited contact with healthcare facilities. These behaviors were investigated by evaluating the incidence of diarrhea and the subsequent healthcare responses, including access to treatment facilities. Weighted averages were employed to analyze healthcare utilization patterns. Given that BHUs play a foundational role in providing accessible healthcare services at the community level, the primary reasons for minimal utilization of BHUs and how they evolved over time (2009-2020) were explored using pareto and rank charts respectively. The Pareto chart effectively illustrated the relative significance of various issues, with an 80% cut-off to highlight the “80/20 rule” [16] which indicates that the most crucial factors, requiring the most attention, typically fall below this threshold. The rank charts were standardized for a population of 10,000 from 2008 to 2020.

#### Regression Analysis

Binomial logistic regression was conducted using pooled data from the 2014– 15 and 2019–20 surveys to examine the association between maternal healthcare utilization and child vaccination status. The primary exposure was maternal engagement with health services (e.g., ANC visits), adjusting for socioeconomic status, maternal education, and geographic factors. The regression model followed the general form:

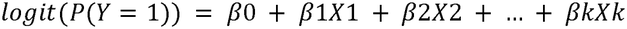

where Y represents zero-dose status (OZD or TZD), and X includes socioeconomic, maternal, and healthcare access variables. Standard errors were adjusted for survey design. Survey weights were applied, and results are reported as adjusted odds ratios to estimate associations; causal interpretations are not inferred due to the cross-sectional nature of the data

### Reporting guidelines

This study is reported in accordance with the Strengthening the Reporting of Observational Studies in Epidemiology (STROBE) guidelines for observational cross-sectional studies. A completed STROBE checklist is provided as Supporting Information (S1 Checklist).

### Ethical Consideration

Ethical approval was not sought for this study because it involved secondary analysis of anonymized, publicly available data from the Pakistan Social and Living Standards Measurement (PSLM) surveys conducted in 2008–09, 2010–11, 2012–13, 2014–15, and 2019–20. No participants were contacted, and the researchers did not access any personally identifiable information. The datasets were used solely for research purposes in accordance with the access and use conditions of the Pakistan Bureau of Statistics.

## Results

Between 2008–09 and 2019–20, the prevalence of operational zero-dose (OZD) children remained low in Punjab, at below 7% in 2008–09, while true zero-dose (TZD) prevalence was below 3.3% in 2010–11. However, there was considerable interprovincial variation. Balochistan exhibited alarmingly high OZD rates of approximately 50% in 2008-09, which slightly decreased to above 35% by 2019-20. In contrast, Sindh (26% to 18%) and KP (18% to 12%) showed improvement over time (Table 2; Figure 1). This temporal improvement, however, was not uniform across the districts.

**Table 2.** Temporal changes in operational and true zero dose prevalence by province (2009-2020)

|  | **2008-09 (%)** | **2010-11 (%)** | **2012-13 (%)** | **2014-15 (%)** | **2019-20 (%)** |
|---|---|---|---|---|---|
| **Punjab** |  |  |  |  |  |
| **Fully Immunized** | 85.2 | 86.1 | 88.7 | 89.4 | 88.8 |
| **Partially Immunized** | 97.0 | 96.7 | 98.2 | 98.8 | 97.1 |
| **OZD** | 6.7 | 6.8 | 4.1 | 3.9 | 5.2 |
| **TZD** | 3 | 3.3 | 1.8 | 1.2 | 1.8 |
| **Sindh** |  |  |  |  |  |
| **Fully Immunized** | 68.9 | 75.3 | 74.1 | 72.9 | 73.2 |
| **Partially Immunized** | 96.9 | 98.5 | 98.2 | 99.2 | 94.4 |
| **OZD** | 25.7 | 20.4 | 20.7 | 17.7 | 18.4 |
| **TZD** | 3.1 | 1.5 | 1.8 | 0.9 | 2.8 |
| **Khyber Pakhtunkhwa** |  |  |  |  |  |
| **Fully Immunized** | 73.6 | 77.5 | 76.1 | 78.2 | 76.9 |
| **Partially Immunized** | 94.7 | 98.3 | 97.7 | 98.4 | 96.9 |
| **OZD** | 18.3 | 16.6 | 17.8 | 16.3 | 11.5 |
| **TZD** | 5.3 | 1.8 | 2.3 | 1.6 | 2.7 |
| **Balochistan** |  |  |  |  |  |
| **Fully Immunized** | 42.9 | 56.7 | 53.3 | 51.1 | 53.2 |
| **Partially Immunized** | 92.8 | 94.5 | 91.8 | 96.1 | 84.7 |
| **OZD** | 49.8 | 42.9 | 42.2 | 45.4 | 36.7 |
| **TZD** | 7.2 | 5.5 | 8.2 | 3.9 | 7.2 |

**Note**: *Fully and partially vaccinated are calculated using basic immunization. The basic immunization includes BCG, PENTA-1, PENTA-2, PENTA-3, Polio-1, Polio-2, Polio-3, and Measles-1 with age bracket from 12 to 23 months. The children who have completed the vaccination schedules are fully vaccinated. Partially vaccinated represents children who received at least one of the vaccines during the schedule.*

**Fig 1.**
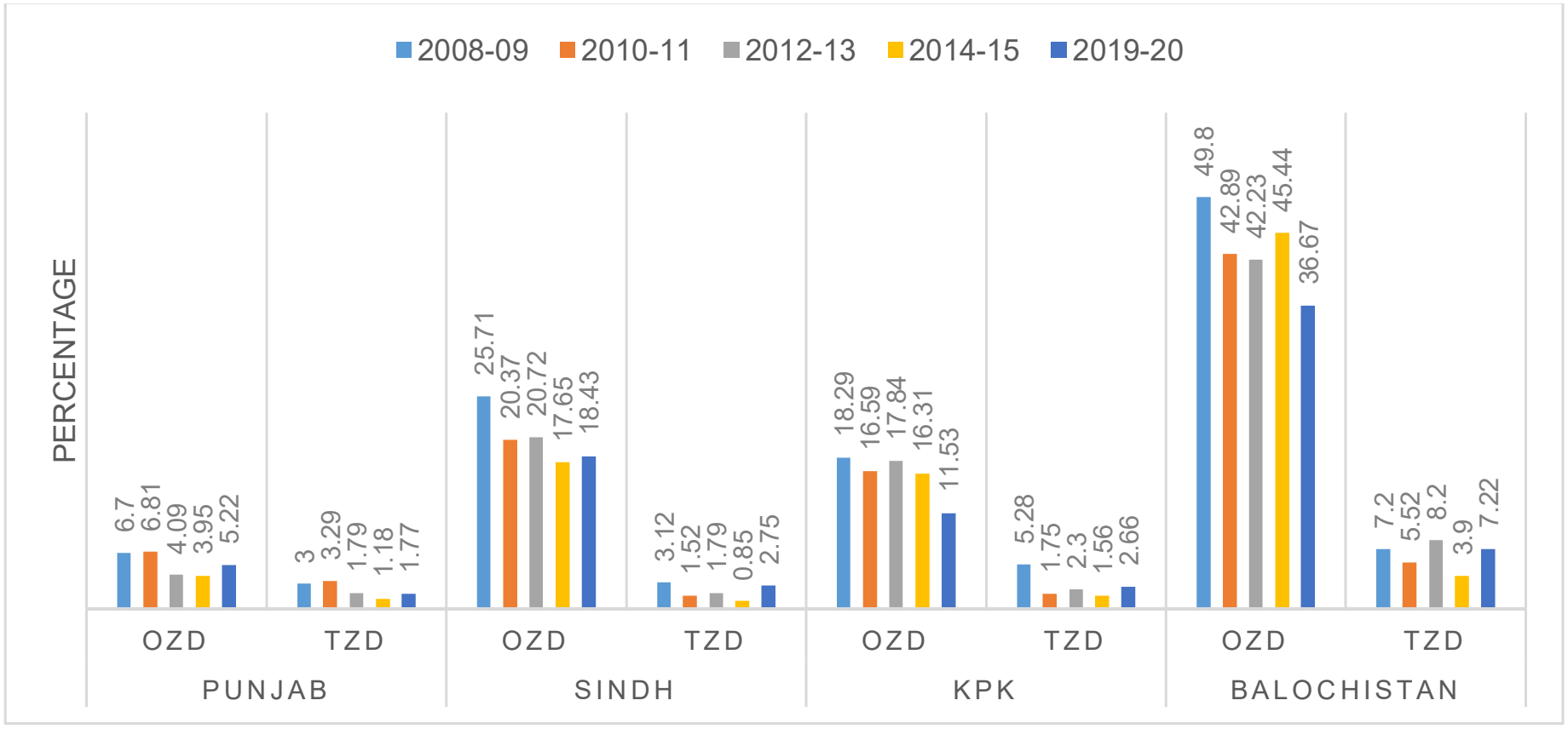
Distribution of operational and true zero dose by province.

Some districts in Sindh, such as Tando Muhammad Khan, Thatta, Tharparkar, and Mirpurkhas, along with various districts in Balochistan like Khuzdar, Washuk, Chagai, Qilla Abdullah, Nasirabad, Jaffarabad, and Kohlu, consistently exhibited higher rates of OZD and TZD. In contrast, Punjab and KP displayed significantly lower rates, with districts in Punjab such as Jehlum, Nankana Sahib, Sialkot, Layyah, and Khushab, along with districts in KP like Malakand, Swabi, Mardan, and Abbottabad, consistently showing the lowest prevalence of zero-dose children.

Fig. 2 presents the spatial distribution of operational and true zero-dose prevalence in relation to overall and healthcare vulnerability between 2008–09 and 2019–20. Punjab shows consistent improvements in all indicators over time, with most districts shifting toward lower vulnerability categories by 2019–20. In contrast, Balochistan, southern Sindh, and parts of northern KP continue to exhibit high levels of overall vulnerability, despite reductions in OZD and TZD in several districts.

**Fig 2.**
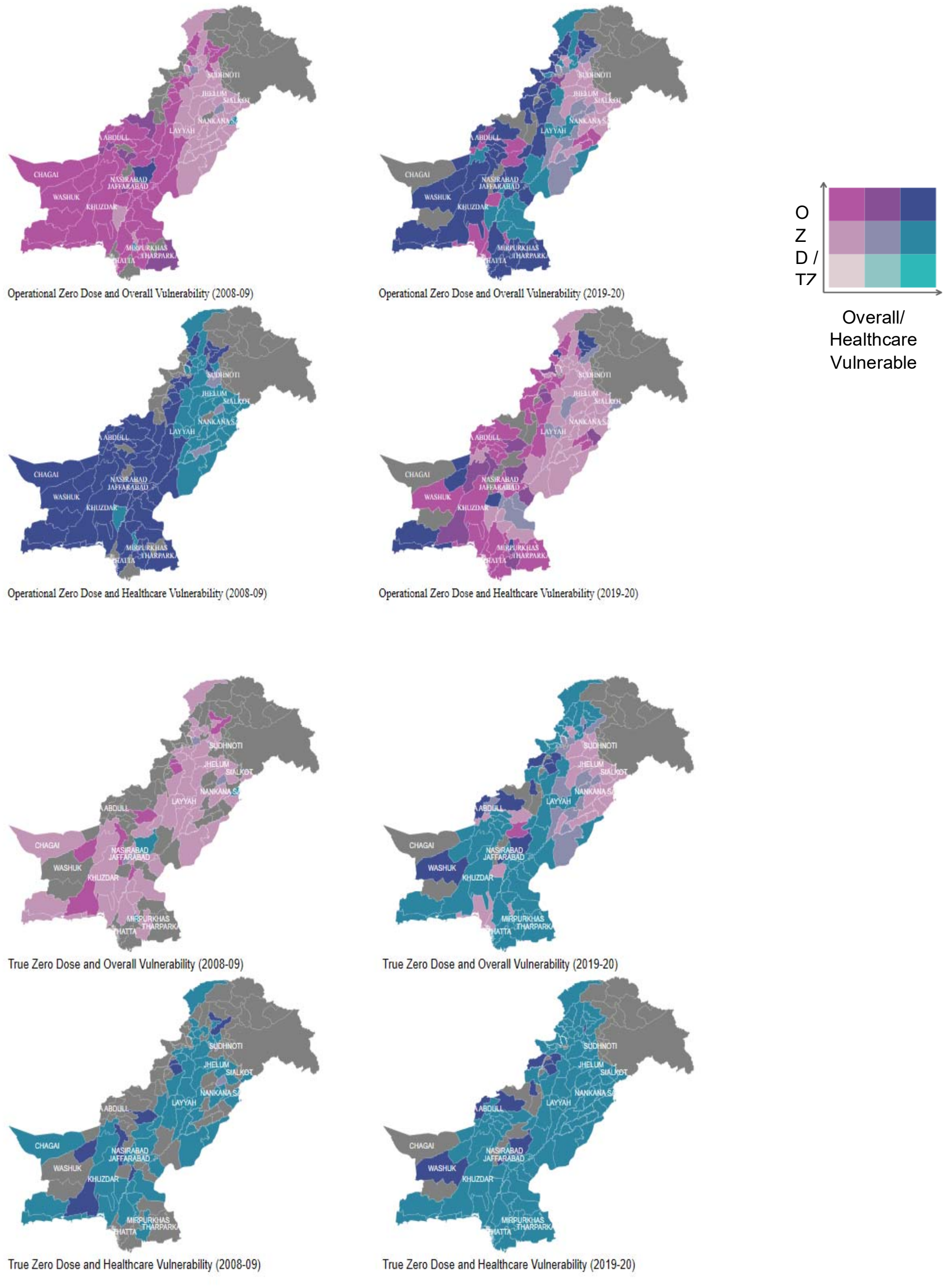
Spatial analysis of operational and true zero doses relative to vulnerability indicators.

For overall vulnerability, the maps indicate an expansion in many districts of Balochistan and parts of Sindh between 2008–09 and 2019–20, reflecting rising socio-economic disadvantage. For healthcare vulnerability, improvements are visible across much of Punjab and KP, while high vulnerability persists in southwestern Balochistan and selected districts of Sindh, mirroring areas with limited health facility availability and weak service readiness (Annex 1).

Fig 3 and 4 presents the reasons for the minimal utilization of healthcare services in districts with high OZD (prevalence ≥50%) and TZD (prevalence ≥10%), particularly in areas like Kohlu, Qilla Abdullah, Qilla Saifullah, Sherani, Washuk (Balochistan), and Torghar and North Waziristan (KP). The primary drivers of non-utilization were distance to health facilities and shortage of health services, while issues such as limited services at the facilities were less frequently cited. These findings were consistent across both the 2012-13 and 2019-20 periods.

**Fig 3.**
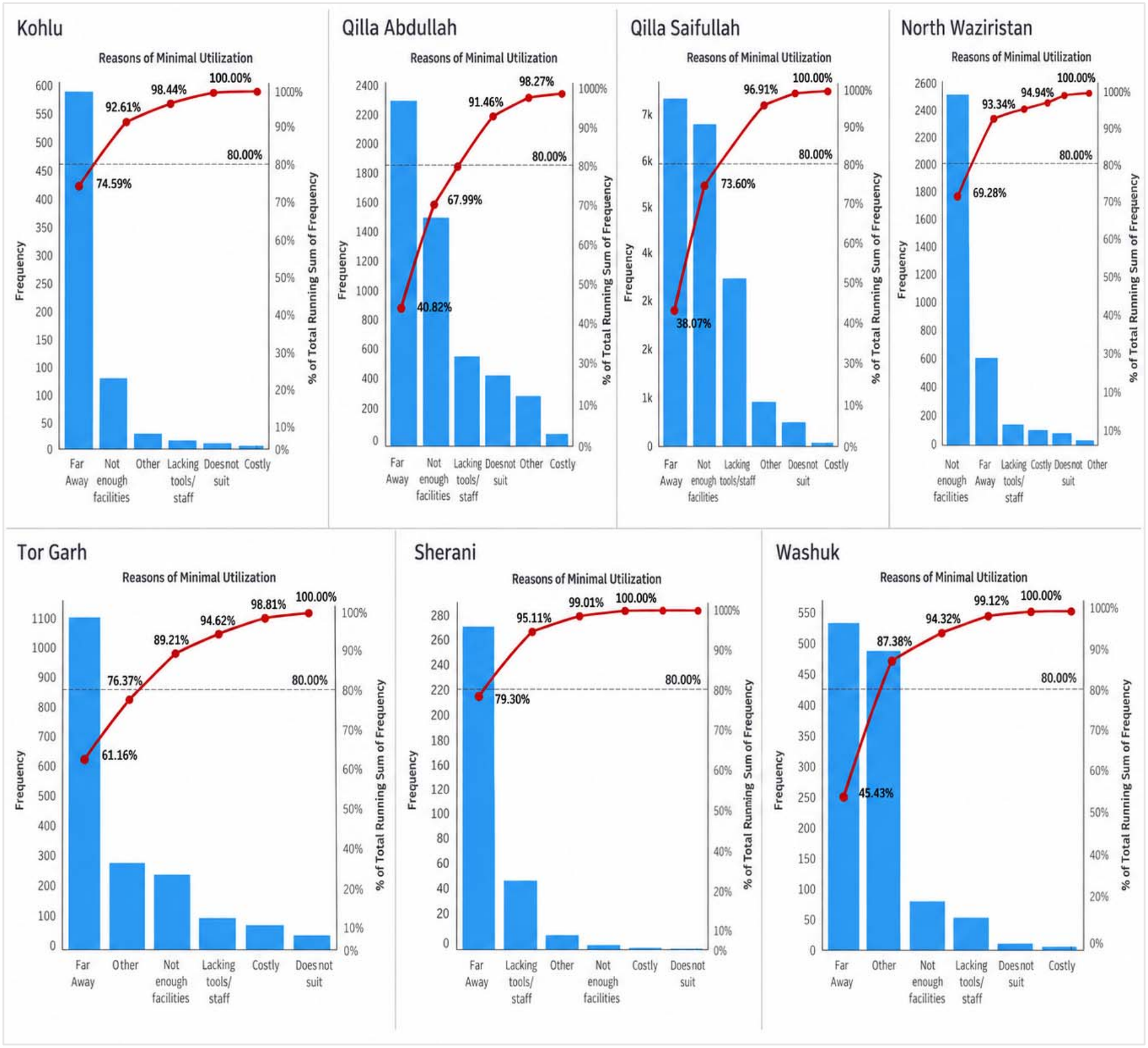
Pareto analysis of reasons for minimal utilization 2019-20. **Note:** *The figure indicates two y-axes; the left axis shows the frequency (calculated from proportions and census data), while the right axis indicates the cumulative percentage*.

**Fig 4.**
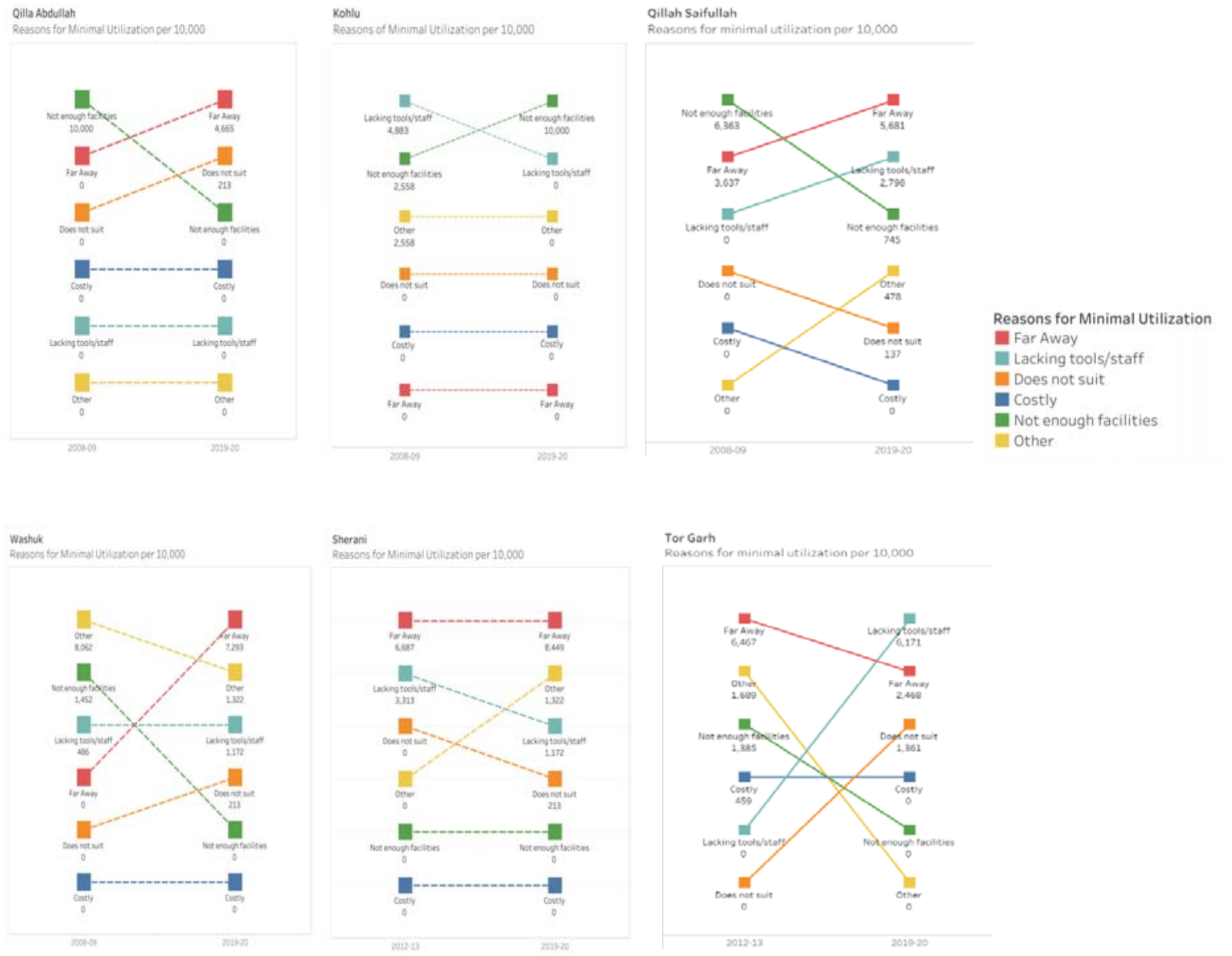
Comparative rank analysis of reasons for minimal utilization for 2008-09 vs. 2019-20.

Table 3 presents multivariable regression estimates examining the association between maternal healthcare experience and zero-dose status among children whose caregivers reported visiting a healthcare facility in the last two weeks. Socioeconomic gradients remain pronounced. Compared to the poorest households, children from middle-income households were 17% less likely to be OZD (OR: 0.829, CI: 0.824–0.834) and 12% less likely to be TZD (OR: 0.876, CI: 0.867–0.886). The effect is stronger for the richest households for OZD (OR: 0.694, CI: 0.689–0.698), while the reduction in TZD is more modest (OR: 0.941, CI: 0.931–0.951).

**Table 3.** Logistic regression of operational and true zero dose prevalence in Balochistan.

| **Covariate** | **OZD, aOR (95% CI)** | **TZD, aOR (95% CI)** |
|---|---|---|
| Wealth Index [Base: Poorest] |  |  |
| Middle | 0.829* | 0.876* |
|  | [0.824,0.834] | [0.867,0.886] |
| Richest | 0.694* | 0.941* |
|  | [0.689,0.698] | [0.931,0.951] |
| Maternal Formal Education [Base: Uneducated] |  |  |
| Primary education | 0.440* | 0.709* |
|  | [0.438,0.443] | [0.702,0.716] |
| Secondary education | 0.275* | 0.489* |
|  | [0.273,0.277] | [0.484,0.494] |
| Higher education | 0.141* | 0.306* |
|  | [0.139,0.143] | [0.300,0.312] |
| Maternal Employment status [Base: Unemployed] |  |  |
| Employed | 1.170* | 1.038* |
|  | [1.166,1.174] | [1.031,1.045] |
| Issues about Healthcare Facility |  |  |
| Doctor not available | 1.869* | 3.341* |
|  | [1.804,1.936] | [3.193,3.495] |
| Staff not cooperative | 2.778* |  |
|  | [2.649,2.913] |  |
| Lack of cleanliness | 8.554* | 30.117* |
|  | [7.704,9.499] | [27.144,33.416] |
| Female staff not available | 0.698* | 0.695* |
|  | [0.651,0.748] | [0.603,0.801] |
| Expensive treatment | 0.906* | 1.249* |
|  | [0.888,0.924] | [1.207,1.292] |
| Year [Base: 2008-09] |  |  |
| 2010-11 | 0.948* | 0.822* |
|  | [0.944,0.953] | [0.816,0.829] |
| 2012-13 | 0.940* | 0.782* |
|  | [0.935,0.946] | [0.773,0.791] |
| 2014-15 | 1.312* | 0.749* |
|  | [1.305,1.320] | [0.740,0.759] |
| 2019-20 | 1.101* | 1.226* |
|  | [1.094,1.107] | [1.213,1.240] |
| Vulnerabilities |  |  |
| Overall vulnerability | 1.304* | 1.099* |
|  | [1.298,1.311] | [1.089,1.110] |
| Healthcare vulnerability | 3.498* | 3.416* |
|  | [3.485,3.512] | [3.392,3.439] |
| Interaction between Provinces and Wealth Index [Base: Punjab # poorest] |  |  |
| Sindh # Middle | 0.788* | 0.549* |
|  | [0.782,0.794] | [0.541,0.556] |
| Sindh # Richest | 0.965* | 0.559* |
|  | [0.957,0.973] | [0.551,0.567] |
| KP # Middle | 2.379* | 0.608* |
|  | [2.360,2.397] | [0.597,0.620] |
| KP # Richest | 1.523* | 0.591* |
|  | [1.508,1.538] | [0.580,0.603] |
| Balochistan # Middle | 5.861* | 1.864* |
|  | [5.805,5.917] | [1.831,1.897] |
| Balochistan # Richest | 3.981* | 0.930* |
|  | [3.935,4.027] | [0.907,0.954] |
| Observations | 77159 | 80656 |

Maternal education shows a strong and consistent gradient. Mothers with primary education were 56% less likely to be OZD and 29% less likely to be TZD compared to children of uneducated mothers. This effect strengthens with higher education, with children of highly educated mothers being 86% less likely to be OZD (OR: 0.141, CI: 0.139–0.143) and 69% less likely to be TZD (OR: 0.306, CI: 0.300– 0.312). In contrast, maternal employment is associated with slightly higher odds of both OZD (OR: 1.170, CI: 1.166–1.174) and TZD (OR: 1.038, CI: 1.031–1.045).

Healthcare facility experiences exhibit strong associations with vaccination outcomes. Children whose caregivers reported doctor unavailability were significantly more likely to be zero-dose, particularly for TZD (OR: 3.341, CI: 3.193–3.495). Similarly, uncooperative staff substantially increased the likelihood of OZD (OR: 2.778, CI: 2.649–2.913), reflecting missed opportunities within the system. Among all variables, lack of cleanliness shows the largest magnitude of association, with children being over eight times more likely to be OZD (OR: 8.554–10.778 across specifications) and up to thirty times more likely to be TZD (OR: 17.937–30.117). Other experience variables show mixed patterns. Reports of female staff unavailability are associated with lower odds of OZD but higher or inconsistent effects on TZD. Similarly, expensive treatment shows a weak or inconsistent association with OZD but increases the likelihood of TZD in some specifications.

Temporal trends indicate significant variation over time. Compared to the 2008–09 baseline, the likelihood of OZD declined in 2010–11 (OR: 0.948, CI: 0.944–0.953) and 2012–13 (OR: 0.940, CI: 0.935–0.946), but increased substantially in 2014–15 (OR: 1.312, CI: 1.305–1.320) and remained elevated in 2019–20 (OR: 1.101, CI: 1.094–1.107). In contrast, TZD declined steadily in earlier rounds, including 2010–11 (OR: 0.822, CI: 0.816–0.829), 2012–13 (OR: 0.782, CI: 0.773–0.791), and 2014–15 (OR: 0.749, CI: 0.740–0.759), before increasing again in 2019–20 (OR: 1.226, CI: 1.213–1.240).

Vulnerability measures show strong and consistent associations. Children residing in households classified as overall vulnerability had higher odds of being OZD (OR: 1.304, CI: 1.298–1.311) and TZD (OR: 1.099, CI: 1.089–1.110). Healthcare-specific vulnerability had an even larger effect, with children whose mothers had low engagement with maternal health services being substantially more likely to be OZD (OR: 3.498, CI: 3.485–3.512) and TZD (OR: 3.416, CI: 3.392–3.439).

Substantial regional disparities persist. Interaction terms reveal that in Balochistan, even children from relatively wealthier households face significantly higher risks of being zero-dose. Compared to the poorest households in the reference province, children in middle-income households in Balochistan were over five times more likely to be OZD (OR: 5.861–6.450), while those in the richest group had nearly four to six times higher odds (OR: 3.981–5.714). For TZD, the effects are smaller but still elevated for middle-income households (OR: 1.728–1.864), while mixed patterns are observed among the richest. In contrast, Sindh shows consistently lower odds (e.g., middle OR: 0.788, TZD OR: 0.549), while Khyber Pakhtunkhwa exhibits a divergence, with higher odds for OZD (OR: 2.379 for middle) but lower odds for TZD (OR: 0.608).

## Discussion

This study shows that Pakistan’s progress in reducing zero-dose prevalence has been real but structurally uneven; national improvements in vaccination coverage have not translated into equitable reach across provinces and districts. Although operational zero-dose prevalence remained above 10% nationally in 2020 and true zero-dose prevalence was below 2.4%, these averages conceal substantial and persistent subnational inequalities. Provincial declines between 2008 and 2020 were sizable_ for example, from 18% to 12% in KP, and from 50% to 37% in Balochistan, yet left the high burden provinces far above Punjab, where prevalence was approximately 5% in 2020. District-level patterns showed that the burden remained concentrated where socioeconomic vulnerability, weak healthcare engagement, and health-system constraints overlapped, rather than being uniformly distributed. These findings suggest that improvements in national vaccination coverage have not translated into equitable reach across provinces and districts.

The distinction between OZD and TZD provides an important perspective on the nature of immunization exclusion. The consistently lower prevalence of TZD suggests that it may identify children experiencing the deepest exclusion from routine immunization services [17]. OZD, by contrast, appears to represent a broader group of children who fail to initiate the routine vaccination schedule because of a combination of household disadvantage, weak contact with maternal and child health services, and operational failures in service delivery [18, 19]. The present analysis cannot fully separate these mechanisms, but the different geographic and socioeconomic patterns associated with the two indicators suggest that they should not be treated as interchangeable. Distinguishing between them may help programs differentiate children who are almost completely disconnected from the health system from those who encounter services but do not successfully initiate routine immunization.

The strong associations with household wealth and maternal education demonstrate that zero-dose status is embedded within broader patterns of social disadvantage. Children from wealthier households had substantially lower odds of being zero-dose (OR 0.5 to 0.8), while higher maternal education was associated with odds ratios of approximately 0.6 to 0.7. Maternal education may influence knowledge of vaccination schedules, confidence in formal healthcare, mobility, and the ability to navigate health services [20, 21]. Household poverty may simultaneously restrict transport, increase the opportunity costs of seeking care, and reduce continuity of contact with maternal and child health services [22]. These findings are consistent with previous evidence from Pakistan showing that immunization gaps disproportionately affect socioeconomically disadvantaged households and children of mothers with limited education [13, 17, 23-26]. They also indicate that zero-dose vaccination is not an isolated immunization problem but one manifestation of wider social and healthcare exclusion.

Service experience emerged as another important determinant of zero-dose status. Long distances to facilities and inadequate availability of services were among the most frequently reported reasons for not using healthcare facilities [27]. Moreover, reported lack of cleanliness showed particularly large associations with zero-dose status (OR 10.8 to 17.9), while uncooperative staff were associated with approximately twice the odds of being zero-dose. These findings should be interpreted cautiously because they are based on reported service conditions and do not establish causality. Nevertheless, their magnitude indicates that the usability and perceived quality of healthcare services may be as important as their geographic availability [28]. Unhygienic environments, poor staff behavior, unavailable providers, limited services, and long waiting times may discourage caregivers from returning to facilities or engaging consistently with routine immunization [29]. Zero-dose prevalence may therefore reflect not only whether a facility exists, but also whether families consider its services accessible, reliable, and acceptable.

This study extends the existing literature on childhood immunization inequity in Pakistan in several ways. Previous research has commonly examined individual determinants of vaccination within selected provinces, districts, or population groups. The present study combines more than a decade of nationally representative data with provincial trends, district-level spatial analysis, vulnerability assessment, and separate examination of operational and true zero-dose indicators. This approach demonstrates that persistent zero-dose burden is geographically clustered rather than uniformly distributed and that similar levels of nominal healthcare access may produce different outcomes depending on the surrounding socioeconomic and service-delivery environment [19, 30]. The analysis therefore moves beyond describing national or provincial coverage gaps by identifying the types of districts in which exclusion persists and the combination of factors that may sustain it.

Differences in provincial administrative and service-delivery capacity may also contribute to the observed geographic inequalities. Following the 18th Constitutional Amendment, responsibility for health-service delivery was transferred more fully to provincial governments. Previous research has suggested that differences in fiscal resources, administrative capacity, and implementation arrangements may influence the consistency of service delivery across provinces [31-33]. However, the present study did not directly assess the effects of decentralization, and the findings should not be interpreted as evidence that decentralization caused provincial disparities. Instead, the results indicate that the same national immunization objectives are being pursued within markedly different provincial and district contexts, requiring strategies that are responsive to local capacity and vulnerability.

The program implications are threefold. First, immunization resources should be geographically targeted toward districts with persistent OZD and TZD burden rather than allocated solely on the basis of provincial averages [19]. Equity-based allocation, updated microplanning, and the use of GIS and vulnerability data could help identify underserved settlements within high-burden districts. Second, increasing the number or proximity of vaccination points should be accompanied by improvements in facility readiness and caregiver experience, including cleanliness, provider availability, staff behaviour, waiting times, and continuity of vaccine services [28, 29]. Third, routine immunization should be more closely connected with maternal and child health contacts and established community outreach platforms. Pakistan’s Polio Eradication Initiative provides an extensive network of frontline workers, community contacts, surveillance systems, and local operational knowledge that could support the identification and referral of zero-dose children [24]. Integration should strengthen routine immunization rather than create parallel delivery arrangements, with particular attention to persistently vulnerable areas in Balochistan and KP. Existing weaknesses in EPI outreach, microplanning, targeting criteria, and data quality will also need to be addressed to prevent programs from repeatedly missing the same communities [34].

Further research is needed to test more directly whether OZD and TZD represent different pathways of exclusion. Future studies should incorporate direct measures of caregiver knowledge, vaccine confidence, gender and mobility restrictions, service trust, conflict-related disruption, and previous experiences with healthcare providers [35]. Finer spatial analysis is also required because district-level estimates may conceal pockets of exclusion in urban informal settlements, remote rural communities, and conflict-affected localities. Prospective and mixed-methods studies could assess whether reported service-quality problems predict subsequent vaccination behavior and clarify why apparently accessible services remain underused. Such evidence would help determine whether interventions should primarily restore first contact with the health system, improve conversion of healthcare contact into vaccination, or address both mechanisms simultaneously.

Overall, the findings indicate that Pakistan’s remaining zero-dose burden is not simply the result of insufficient geographic access. It is concentrated where socioeconomic disadvantage, weak maternal-health engagement, and poor service experience intersect. Sustained and equitable progress will therefore require geographically targeted strategies that improve both the reach and the quality of routine immunization while addressing the broader conditions that prevent marginalized families from using available services.

## Limitations

This study has several limitations. The analysis relied on secondary PSLM data, and vaccination status was determined using caregiver recall and available vaccination cards, creating the possibility of recall and classification bias. The absence of comparable survey data between 2015 and 2019 limited assessment of short-term changes in immunization coverage. Available variables did not permit detailed examination of cultural norms, vaccine attitudes, gender-related mobility restrictions, insecurity, conflict-related disruptions, or other contextual influences on vaccination. Reported reasons for healthcare non-use and service-quality problems may also reflect caregiver perceptions and cannot be interpreted as objective facility assessments.

District-level aggregation may conceal substantial within-district disparities, particularly in urban informal settlements and geographically remote rural communities. Although the operational and true zero-dose measures provide complementary indicators of exclusion, they may not capture all partial, undocumented, or informal vaccination occurring outside the routine EPI reporting framework. The cross-sectional design precludes causal inference, and residual confounding from unmeasured household, community, and health-system characteristics remains possible. In addition, the thresholds used to identify benchmark and high-risk districts may not fully capture differences in local context. Nevertheless, the consistency of the temporal, spatial, and regression findings suggests that these limitations are unlikely to fully account for the observed inequalities.

## Conclusion

Pakistan has achieved meaningful reductions in zero-dose prevalence, but these gains have not been distributed equitably. Persistent OZD and TZD burden remains concentrated in Balochistan, parts of KP and Sindh, and other districts where socioeconomic vulnerability, weak maternal-health engagement, and health-service constraints overlap. Household poverty and low maternal education remain important predictors, but the strong associations with cleanliness, staff behaviour, and service availability also show that nominal access does not necessarily translate into acceptable or effective care.

Closing the remaining immunization gap will therefore require more than expanding the physical availability of vaccination services. Pakistan needs equity-sensitive district targeting, updated and spatially informed microplanning, improved frontline service quality, stronger links with maternal and child health services, and careful use of PEI outreach infrastructure to strengthen routine immunization. Without attention to both social exclusion and service experience, improvements in national coverage may continue to coexist with persistent pockets of zero-dose children.

## Data Availability

No new data were generated for this study. The analyses used existing, anonymized data from the Pakistan Social and Living Standards Measurement (PSLM) surveys for 2008-09, 2010-11, 2012-13, 2014-15, and 2019-20. These datasets are available from the Pakistan Bureau of Statistics (PBS) through its official data access procedures and website. The authors had no special access privileges to the data that other researchers would not have.

## ANNEXURE 1: Bivariate Maps of Zero Dose Status and Vulnerabilities (2010-2015)

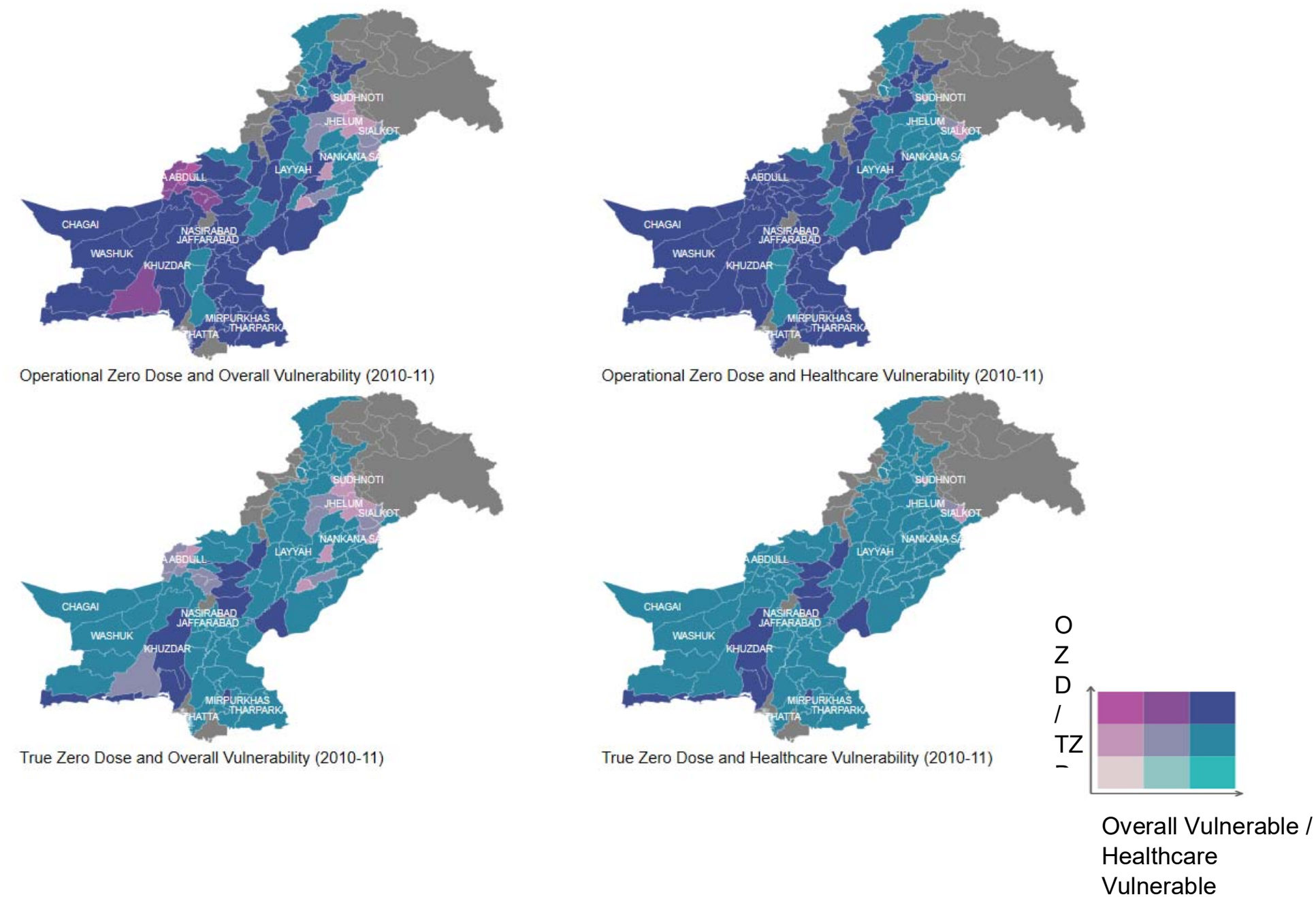
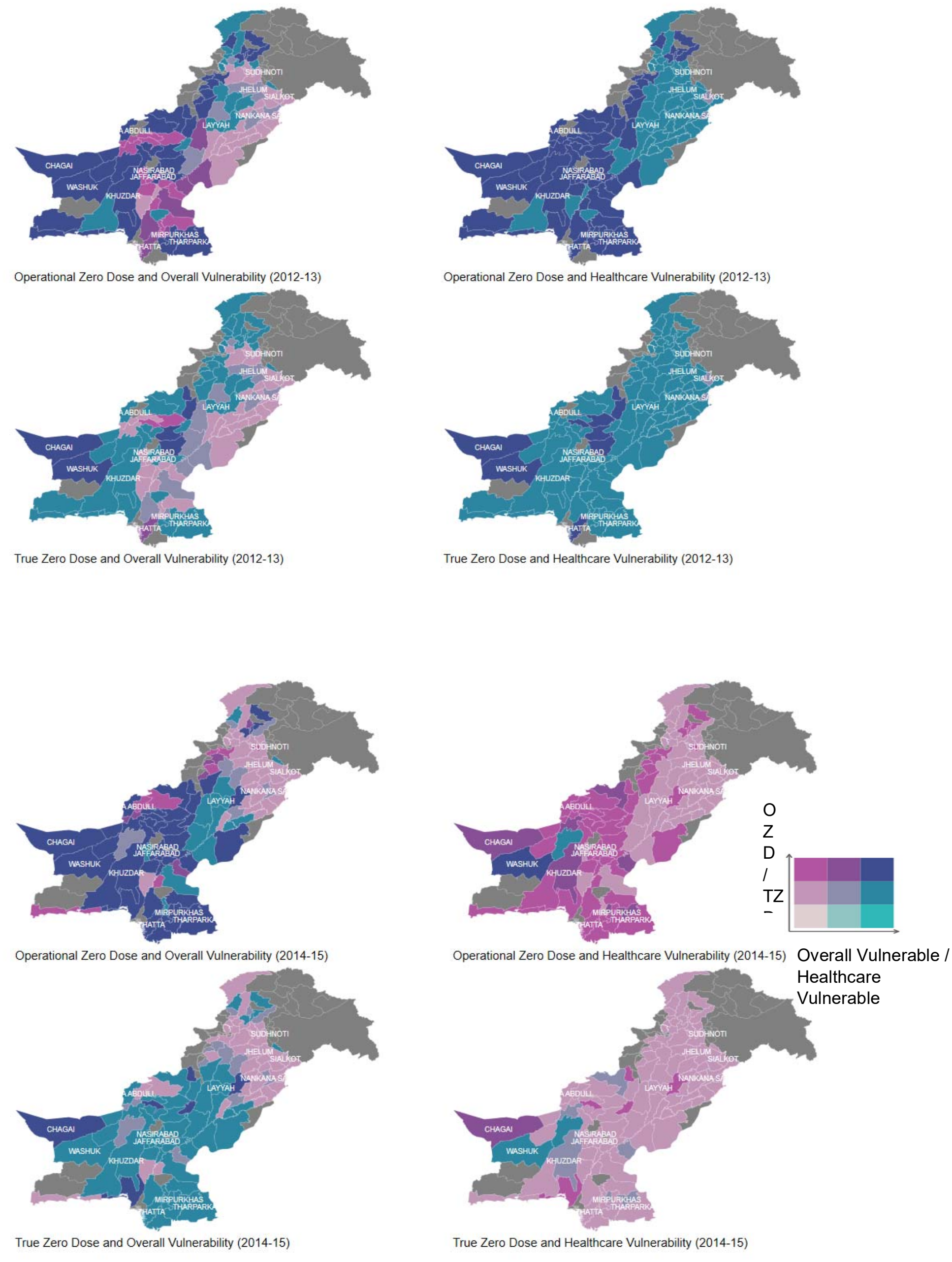

## Supporting information

S1 Checklist. STROBE checklist for cross-sectional studies.

